# Tuberculosis Severity metrics poorly predict Two-Month Culture Conversion

**DOI:** 10.64898/2026.11.19.26368488

**Authors:** Lentlametse Mantshonyane, Penelope Benchek, LaShaunda Malone, Mary Nsereko, Hussein Kisingo, Daniel J. Tisch, Fredrick R. Schumacher, W. Henry Boom, Harriet Mayanja-Kizza, Catherine M. Stein, Scott M. Williams

**Affiliations:** Department of Population and Quantitative Health Sciences, Case Western Reserve University, Cleveland, OH; Department of Medicine, Division of Infectious Diseases, Case Western Reserve University, Cleveland, OH; Uganda-CWRU Research Collaboration, Kampala, Uganda; School for Biomedical Sciences, College of Health Sciences, Makerere University; Department of Medicine, School of Medicine, Makerere University and Mulago Hospital, Kampala Uganda; Cleveland Institute for Computational Biology, Cleveland, OH

**Keywords:** Tuberculosis outcomes, Mycobacterium clearance, TB severity scores

## Abstract

**Objectives:** Among people with pulmonary tuberculosis (PTB), culture conversion (bacterial clearance) after two months of treatment is a critical indicator of response. However, predictors of culture conversion at two months are difficult to define and can be context specific. In this study, we evaluated how well two tuberculosis (TB) severity metrics, the chest Xray (CXR) Score and the Bandim TBScore associate with and predict two-month culture conversion.

**Methods:** The cohort contained 718 index PTB cases enrolled in “The Kawempe Community Health Study (KCHS)”. We analyzed severity metrics, demographics, clinical, microbiological, radiological, household and behavioral data for their association with and their ability to predict culture conversion.

**Results:** Sixty-eight percent of participants culture converted at two months. The highest CXR Score category (odds ratio (OR) 0.51, 95% Confidence Interval (CI) 0.31, 0.84), smear grade at diagnosis (OR 0.42, 95% CI; 0.26, 0.62), pleural thickening (OR 0.41, 95% CI 0.17, 0.95) and current smoking (OR 0.31, 95% CI 0.09, 0.99) inversely associated with conversion. Both severity measures and the most parsimonious model poorly predicted culture conversion (all AUCs<0.70).

**Conclusion:** CXR Score, higher smear grade at diagnosis, pleural thickening, and current smoking inversely associated with culture conversion but provided limited utility for prediction.

---

Pulmonary tuberculosis (PTB) disease is the leading infectious cause of death worldwide, with an estimated 1.26 million deaths in 2024[1]. PTB is caused by the bacterium *Mycobacterium tuberculosis* (Mtb)[1]. Although most people infected with Mtb do not develop active PTB, in those who do, variation exists in the disease course, severity, and treatment outcomes. One of the most important treatment and programmatic goals of PTB care is to eliminate the Mtb from the sputum of a patient within the first two months of treatment, often referred to as two-month culture conversion. From a clinical perspective, failure to convert cultures to negative after two months indicates poor treatment response and is positively correlated with several adverse outcomes[2], including mortality[3], development of drug resistance, prolonged duration of treatment, and sequelae such as post-tuberculous lung disease (PTLD)[4, 5]. From a public health perspective, individuals who do not convert can continue to infect other people, as an untreated PTB patient is estimated to infect about 10-15 individuals annually[6, 7].

Two-month culture conversion is difficult to predict[4] but previous studies of drug sensitive PTB have found associations with baseline smear grade[8, 9], medication adherence[8, 9], radiographic pulmonary lesions[8], HIV positive status[9, 10]. In multi-drug resistant TB (MDR-TB) co-morbidities[2], low body mass index (BMI), drug resistance profile and medication adherence were identified as associating with two-month conversion [6]. Nonetheless, predictors of conversion have proven difficult to understand as they can vary by context including clinical settings, population and drug resistance profile[5, 6, 11, 12]. Another potential predictor of culture conversion is the severity of PTB at diagnosis, an important trait that impacts patients’ experience of PTB, quality of life, risk for long term sequelae[5, 13] and mortality[14]. However, it is unknown how disease severity associates with or predicts two-month culture conversion.

Two of the most used severity grading metrics in different settings are the Bandim TBScore[8, 14–17] and a composite chest X-ray (CXR) score[8]. The Bandim TBScore is a composite metric used to grade PTB severity based on the presence of five self-reported symptoms and six observable signs[18]. Each sign and symptom contributes one point while BMI and mid upper arm circumference can each contribute an extra point if they are extreme (<16kg/m2 or <200mm, respectively) for a maximum of 13 points[14, 18]. It was developed and validated for use in resource limited settings and predicts other outcomes such as mortality[14, 19, 20].

The CXR is also widely used for severity grading[21] and treatment evaluation[22, 23]. Severe PTB on CXR is reflected by the presence of radiographic abnormalities [16] (Supplemental Table 1). A simple, numerical CXR Score derived from the estimated proportion of lung affected and presence of cavitations was developed and validated in Indonesia[8, 24]. CXR Score is calculated by the formula: *CXR Score =* percentage of lung affected plus 40 if cavitations are present[8].

If these cost-effective and accessible severity metrics have good predictive utility for conversion, they can be used to stratify patients unlikely to culture convert to prioritize them for intensified targeted early interventions that enhance conversion. Identifying predictors of conversion might also inform the design of setting specific intervention strategies.

In this study, we examined how well the two composite severity measures, CXR Score and Bandim TBScore and other routinely collected demographic, microbiology, household, health behaviors and co-morbidity variables associate with or predict culture conversion in urban Uganda.

## Methods

### Study Population and Materials

Data for this cohort study were from the “Kawempe Community Health Study (KCHS)”, a household contact study in which 872 culture-confirmed PTB cases and 2,585 household contacts were enrolled from 2002–2012 in Kampala, Uganda[25]. The parent study excluded individuals with relapse or prior PTB. Of the 872 adult (over the age of 18 years) index cases, 718 had complete culture conversion data and were included in the subsequent analyses. All cases had drug sensitive Mtb. We excluded household contacts because shared genetic background (genetic relatedness) may affect both disease severity[15] and conversion[26]. We defined culture conversion as negative Mtb culture in liquid media test after two months of treatment in participants initially diagnosed as Mtb positive from sputum. Comparisons of baseline characteristics between included and excluded individuals are presented in Supplementary Table 1.

#### Human subjects

The study was approved by the National HIV/AIDS Research Committee of Makerere University and the institutional review board at University Hospitals, Cleveland Medical Center. Final clearance was given by the Uganda National Council for Science and Technology. Written informed consent was obtained from all study participants.

#### Study exposures

Our primary analyses assessed the associations between the two severity grading metrics: the Bandim TBScore and CXR Score as independent variables and culture conversion at two months as the dependent variable. Unadjusted and sex, age and HIV adjusted logistic regression models were used to assess these associations.

Bandim TBScore was calculated as adapted and described previously for Uganda[27]. The CXR Score was estimated using the formula: CXR Score = (number of affected lung zones / 6) *100 plus 40 if cavitations are present[8]. This score was not normally distributed; therefore, we converted it into quartile categories.

#### Statistical Analyses Association

To explore whether individual components of the Bandim TBScore or CXR measures associate with culture conversion better than the composite scores, we performed univariate association analyses for each component of the Bandim TBScore and each CXR measure with conversion. Independent variables were modeled as continuous, categorial or ordinal, where appropriate. Significance for single variable association was defined as p ≤ 0.05. We then built multivariate models including Bandim TBScore components or CXR measures that were significant in the univariate analysis.

#### Variable Selection LASSO Model

Least Absolute Shrinkage and Selection Operator (LASSO) logistic regression[28] (glmnet, alpha = 1) was fit to the variables significant in univariate screening (p < 0.10), some of which are correlated (r^2^ > 0.5)(Supplementary Tables 2). Predictors were standardized, and ordinal predictors were treated as numeric in the model. The penalty parameter λ was chosen by 10-fold cross-validation using AUC as the criterion, selecting the largest λ within one standard error of the maximum CV AUC. A random seed was used for replication.

LASSO selected variables were re-fit using multivariate regression to evaluate their unpenalized effects. All multivariate regression models were adjusted for age, sex, and HIV status because they are known confounders. Predictors were assessed for multicollinearity, using variance inflation factors (VIFs) [28–30]. Our analyses showed no evidence of significant multicollinearity (VIFs << 5).

#### Prediction Models

Prediction of conversion, using the area under the receiver operating (AUC) characteristics, were then performed with different combinations of variables.

#### Dominance Analysis

We assessed the relative importance of the selected features most associated with culture conversion using dominance analysis (DA)[31]. In a logistic regression model, DA evaluates the importance of each predictor in the model based on how much it contributes to the prediction of the outcome than does its competitor predictor on average (general dominance) and when it is added at a given level of analysis (conditional dominance)[31, 32]. General dominance indicates how the overall fit statistic measure is divided among included features, while conditional dominance assesses the measure associated with a feature based on the position (level) at which it is included in the model[31, 32]. While we assessed the relative contributions using multiple measures, we reported the McFadden R-square as it meets all four criteria for best measures of feature importance[32].

All missing data for independent variables were imputed using predictive mean matching[33]. Significance level was set at p<0.05. All analyses were done in R programming version 4.4.1 (2024-06-14).

## Results

Seven hundred and eighteen (718) of the 872-index adult PTB cases who had data on two-month culture conversion contributed to these analyses. Participants excluded from the analysis for not having data on culture conversion were similar to those included in most characteristics with the following exceptions. They were more likely to be anemic (48% versus 12%, p<0.001), were thinner (have low fat mass index (4.29 kg/m^2^ versus 5.40 kg/m^2^, p=0.007), have disease exclusively affecting the left lung (52% versus 22%, p<0.001), have pleural effusion (13% versus 6.7%, p=0.01) and were more likely to die (14.6% versus 4.5%, p<0.001) than those included in our study (Supplementary Table 1).

The distributions of baseline characteristics univariately associated with two-month culture conversion indicated that most included participants (68%) culture converted. People who culture converted were more likely to be HIV positive, to have lower smear grade, to have fewer affected lung zones, to have never smoked tobacco, and to live in houses with fewer windows (Table 1). Other associated variables included: muzingo type of house, more people in household, pleural thickening, lung infiltrates, lung fibrosis, being admitted to hospital, having higher disease extent and CXR Score (Table 1). Other baseline characteristics and their distribution by culture conversion showed no evidence of association (Supplementary Table 3).

**Table 1.** The distribution of characteristics associated with culture conversion at p-value threshold of p≤0.10 in the cohort.

| Characteristics | N (%)<br>Mean (SD) | Culture conversion at two-months<br>N (%), Mean (SD) |  | P value |
|---|---|---|---|---|
|  |  | Yes | No |  |
|  | N=718 | 489 (68) | 229 (32) |  |
| **Demographic/Clinical features** |  |  |  |  |
| Age in years (mean (SD)) | 29.43 (9.04) | 29.11 (8.69) | 29.58 (9.51) | 0.510 |
| Sex: Male | 376 (52%) | 245 (50.1) | 131 (57.2) | 0.090 |
| HIV status: Positive | 210 (29%) | 161 (32.9) | 49 (21.4) | 0.002 |
| Smear grade (%) |  |  |  | <0.001 |
| Scanty | 22 (3.1%) | 21 (4.3) | 1 (0.4) |  |
| 1+ | 21 (2.9%) | 21 (4.3) | 0 (0.0) |  |
| 2+ | 85 (11.8%) | 67 (13.7) | 18 (7.9) |  |
| 3+ | 590 (82.2%) | 380 (77.7) | 210 (91.7) |  |
| Tobacco use (%) |  |  |  | <0.001 |
| Never smoked | 589 (82%) | 414(84.7) | 175 (76.4) |  |
| Former smoker | 116 (16%) | 70 (14.3) | 46 (20.1) |  |
| Currently smoke | 13 (1.8%) | 5 (1.0) | 8 (3.5) |  |
| Admitted (%) | 72 (10%) | 60 (12.3) | 12 (5.2) | 0.005 |
| **Household features** |  |  |  |  |
| Type of house: Muzigo (%) | 506(70%) | 332 (67.9) | 174 (76.0) | 0.033 |
| People per house (median (IQR) | 4 (3, 6) | 4(3,6) | 4(3,5) | 0.038 |
| Number of rooms (mean (SD)) | 2.21 (1.91) | 2.30 (1.92) | 2.02 (1.87) | 0.068 |
| No. windows (mean (SD)) | 1.41 (1.65) | 1.51 (1.75) | 1.21 (1.39) | 0.022 |
| Type of wall: mud/wattle (%) | 191 (27%) | 120(25) | 71(31) | 0.071 |
| **BandimTBScore measures** |  |  |  |  |
| Tachycardia(pulse>90 beats/min) | 561 (78%) | 392 (80.2) | 169 (73.8) | 0.068 |
| **Chest X-ray measures** |  |  |  |  |
| CXR score (mean (SD)) | 77.69 (37.62) | 74.40 (38.30) | 84.72 (35.21) | <0.001 |
| Disease extent (%) |  |  |  | 0.004 |
| Normal | 30 (4.2%) | 26 (5.3) | 4 (1.7) |  |
| Minimal | 58 (8.1%) | 47 (9.6) | 11 (4.8) |  |
| Moderate | 250 (35%) | 175 (35.8) | 75 (32.8) |  |
| Advanced | 380 (53%) | 241 (49.3) | 139 (60.7) |  |
| Affected Lung |  |  |  | 0.035 |
| Both lungs | 361(50%) | 235 (48.1) | 126 (55.0) |  |
| Right lung | 196 (27%) | 133 (27.2) | 63 (27.5) |  |
| Left lung | 161 (22%) | 121 (24.7) | 40 (17.5) |  |
| No. affected lung zones (mean (SD)) | 3.12 (1.64) | 2.97 (1.64) | 3.46 (1.60) | <0.001 |
| Lung fibrosis (%) | 82 (11%) | 47 (9.6) | 35 (15.3) | 0.036 |
| Infiltrated lobe (%) |  |  |  | 0.014 |
| None | 29 (4.0%) | 27 (5.5) | 2 (0.9) |  |
| Lower lobe | 75 (10%) | 53 (10.8) | 22 (9.6) |  |
| Upper lobe | 112 (16%) | 80 (16.4) | 32 (14.0) |  |
| Both lobes | 502 (70%) | 329 (67.3) | 173 (75.5) |  |
| Pleural thickening (%) | 26 (3.6%) | 11 (2.2) | 15 (6.6) | 0.008 |

In unadjusted univariate analyses, participants in the two upper CXR Score quartiles had lower odds of culture conversion than those in the lowest quartile ( **Table *2***a). However, in the analysis, adjusted for age, sex and HIV status, only participants with the most severe PTB, those in the highest CXR Score (q4) category had lower odds of culture conversion (OR 0.51, 95% CI 0.31, 0.84) (Table 2a). The Bandim TBScore was not associated with two-month culture conversion in either unadjusted or adjusted analysis (Table 2a).

**Table 2a.** The unadjusted and the age, sex and HIV adjusted associations between composite severity scores and culture conversion.

| **Chest X-ray Score** | **Unadjusted** |  | **Adjusted** |  |
|---|---|---|---|---|
|  | **OR (95% C I)** | **p-value** | **OR (95% C I)** | **P value** |
| CXR score: *referent Q1(Lowest)* |  |  |  |  |
| *Q2(Lower-Mid)* | 0.64 (0.40, 1.01) | 0.06 | 0.70 (0.44, 1.12) | 0.14 |
| *Q3(Upper-Mid)* | 0.64 (0.42, 0.97) | 0.03 | 0.72 (0.47, 1.10) | 0.14 |
| *Q4(Highest)* | 0.47 (0.28, 076) | <0.01 | 0.51 (0.31, 0.84) | 0.01 |
| Age | 0.99 (0.98, 1.01) | 0.51 | 0.99 (0.97, 1.01) | 0.24 |
| Sex: Male | 0.75 (0.55, 1.03) | 0.08 | 0.85 (0.61, 1.19) | 0.35 |
| HIV status: Positive | 1.80 (1.26, 2.62) | <0.01 | 1.75 (1.20, 2.58) | <0.01 |
| **Bandim TBScore** | **Unadjusted** | **Adjusted** |  |  |
|  |  | **p-value** | **OR (95% C I)** | **p.value** |
| Bandim TBScore | 1.00 (0.92, 1.07) | 0.93 | 0.98 (0.91, 1.06) | 0.69 |
| Age | 0.99 (0.98, 1.01) | 0.51 | 0.99 (0.97, 1.01) | 0.28 |
| Sex: Male | 0.75 (0.55, 1.03) | 0.08 | 0.81 (0.59, 1.13) | 0.21 |
| HIV status: Positive | 1.80 (1.26, 2.62) | <0.01 | 1.85 (1.27, 2.72) | <0.01 |

**2b.** The Unadjusted and the age, sex and HIV adjusted associations between the Chest X-ray features associated with culture conversion.

| **Chest X-ray measures** | **Unadjusted OR (95% CI)** | **p-value** | **Adjusted OR (95% CI)** | P **value** |
|---|---|---|---|---|
| Disease extent: referent: Normal |  |  |  |  |
| Minimal | 0.66 (0.17, 2.14) | 0.51 | 1.51 (0.31, 6.76) | 0.59 |
| Moderate | 0.36 (0.10, 0.96) | 0.06 | 0.92 (0.19, 3.87) | 0.91 |
| Advanced | 0.27 (0.08, 0.70) | 0.02 | 0.89 (0.17, 4.04) | 0.88 |
| No. affected lung zones | 0.84 (0.76, 0.92) | <0.01 | 0.82 (0.68, 0.99) | 0.04 |
| Pleural thickening | 0.33 (0.14, 0.72) | 0.01 | 0.39 (0.16, 0.91) | 0.03 |
| Affected lung: ref: both | 1.0 |  |  |  |
| Right lung | 1.05 (0.72, 1.52) | 0.81 | 0.56 (0.32, 0.95) | 0.03 |
| Left lung | 1.61 (1.06, 2.47) | 0.03 | 0.92 (0.52, 1.63) | 0.77 |
| Infiltrates: ref: None | 1.0 |  |  |  |
| Lower lobe | 0.27 (0.06, 0.86) | 0.05 | 0.31 (0.06, 1.30) | 0.13 |
| Upper lobe | 0.28 (0.06, 0.86) | 0.05 | 0.47 (0.08, 2.10) | 0.36 |
| Both lobes | 0.21 (0.05, 0.61) | 0.01 | 0.46 (0.08, 1.98) | 0.33 |
| Lung fibrosis: yes | 0.59 (0.37, 0.95) | 0.03 | 0.78 (0.47, 1.32) | 0.35 |
| Pleural effusion: yes | 0.76 (0.41, 1.44) | 0.38 |  |  |
| Miliary disease: yes | 1.21 (0.57, 2.79) | 0.64 |  |  |
| Lung adenopathy: yes | 2.00 (0.91, 5.03) | 0.11 |  |  |
| Lung cavities: yes | 0.77 (0.55, 1.07) | 0.13 |  |  |
| Age | 0.99 (0.98, 1.01) | 0.51 | 0.99 (0.97, 1.01) | 0.39 |
| Sex: male | 0.75 (0.55, 1.03) | 0.08 | 0.90 (0.64, 1.27) | 0.56 |
| HIV status: positive | 1.80 (1.26, 2.62) | <0.01 | 1.71 (1.15, 2.57) | 0.01 |

Several individual CXR measures were univariately associated with conversion (Table 2b) while no component of the Bandim TBScore was associated (Supplementary Table 4). After adjusting the model for age, sex and HIV status, pleural thickening (OR 0.39, 95% CI 0.16, 0.91), number of affected lung zones (OR 0.82, 95% CI 0.68, 0.99) and disease exclusively affecting the right lung versus both lungs (OR 0.56, 95% CI 0.32, 0.95) associated with lower odds of conversion (Table 2b).

LASSO was then applied to variables from Table 1 that were significant in the univariate analyses (p<0.1) to select the best associating model with culture conversion. Nine features were selected: baseline smear grade, pleural thickening, hospital admission, tobacco smoking, disease extent on CXR, number of affected lung zones, number of windows in the house, type of house wall, and HIV status. Of these, higher baseline smear grade (OR 0.42, 95% CI 0.26, 0.62), pleural thickening (OR 0.41, 95% CI 0.17, 0.95) and current tobacco smoking versus never smokers (0.31, 95% CI 0.09, 0.99) independently associated with lower odds of culture conversion, while HIV positivity associated with higher odds (OR 1.56, 95% CI 1.04, 2.36) (Table 3).

**Table 3:**
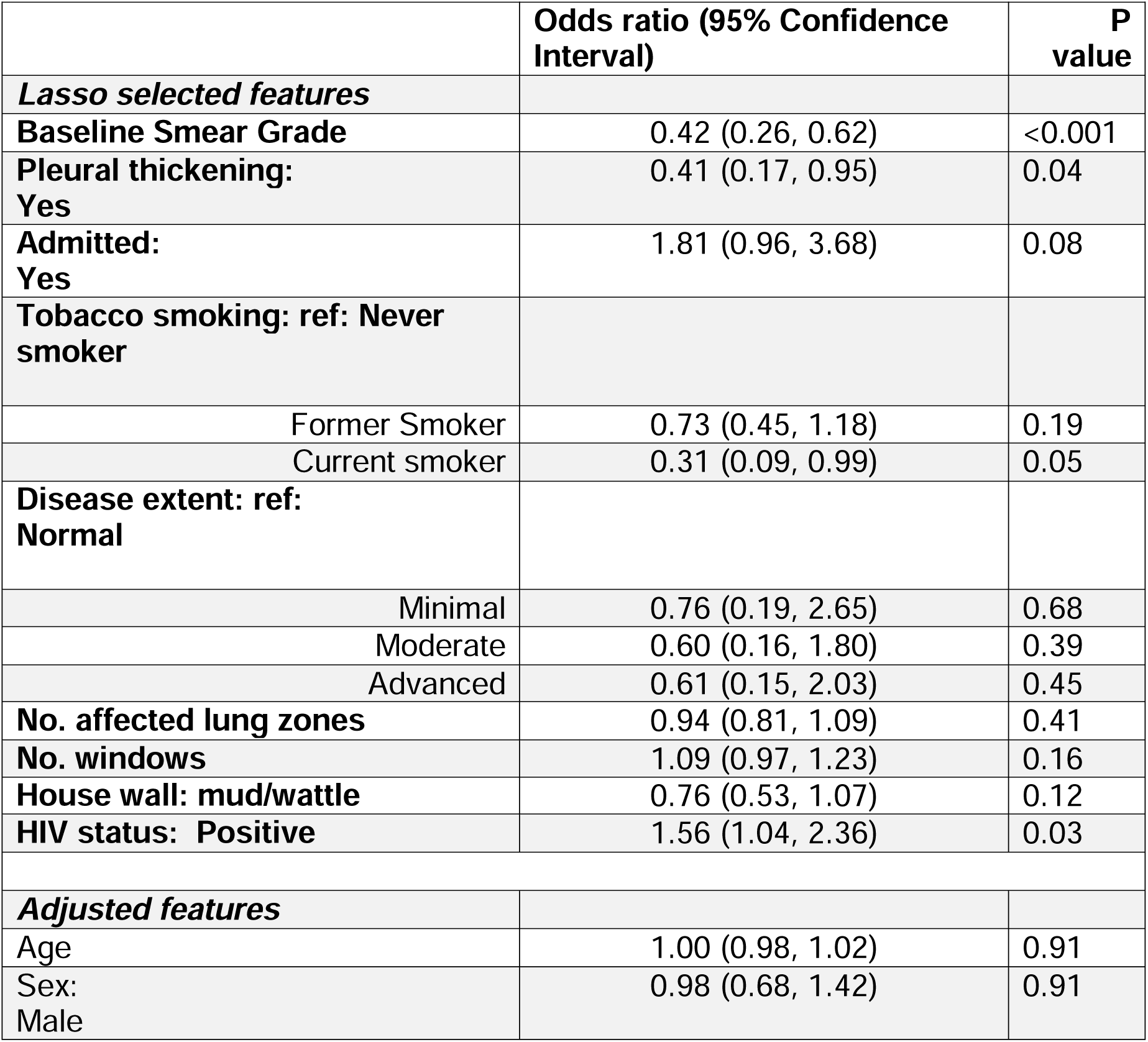
LASSO selected features associated with two-month culture conversion (most parsimonious model)

The age, sex, and HIV adjusted models with Bandim TBScore and with CXR Score both poorly predicted culture conversion with AUCs of (0.57, 95% CI 0.55, 0.64) and (0.59, 95% CI 0.57, 0.65), respectively (Table 4). Combining these composite severity scores into a single age, sex and HIV adjusted model did not improve the prediction of culture conversion. The model with all associated CXR measures, adjusted for age, sex and HIV predicted culture conversion better than models with either composite score. However, it was still not very predictive (AUC 0.62, 95% CI 0.59, 0.68) (Table 4). Including all nine features from the LASSO selection above improved culture conversion prediction but it was still a poor predictor (AUC 0.67, 95% CI 0.63, 0.71) (Table 4).

**Table 4:** Fit statistics and predictive ability (AUC) of all age, sex and HIV adjusted models showing limited prediction.

| Model | AIC | BIC | AUC |
|---|---|---|---|
| Bandim TBScore | 895.2 | 918.1 | 0.57 |
| CXR score | 891.8 | 923.9 | 0.59 |
| CXR score + Bandim TBScore | 894.2 | 930.8 | 0.59 |
| Associated CXR Features only | 882.7 | 923.9 | 0.62 |
| LASSO selected features only | 859.0 | 941.4 | 0.67 |

Dominance analysis was done to evaluate the relative contributions of the LASSO selected features associated with culture conversion. In both the general (Figure 1a) and conditional dominance analyses (Figure 1b), baseline smear grade was the most important feature. HIV status, tobacco smoking and disease extent were only conditionally dominant (Figure Legends Figure *1*b). Other measures of dominance produced similar conclusions (Supplementary Table 5).

**Figure 1.**
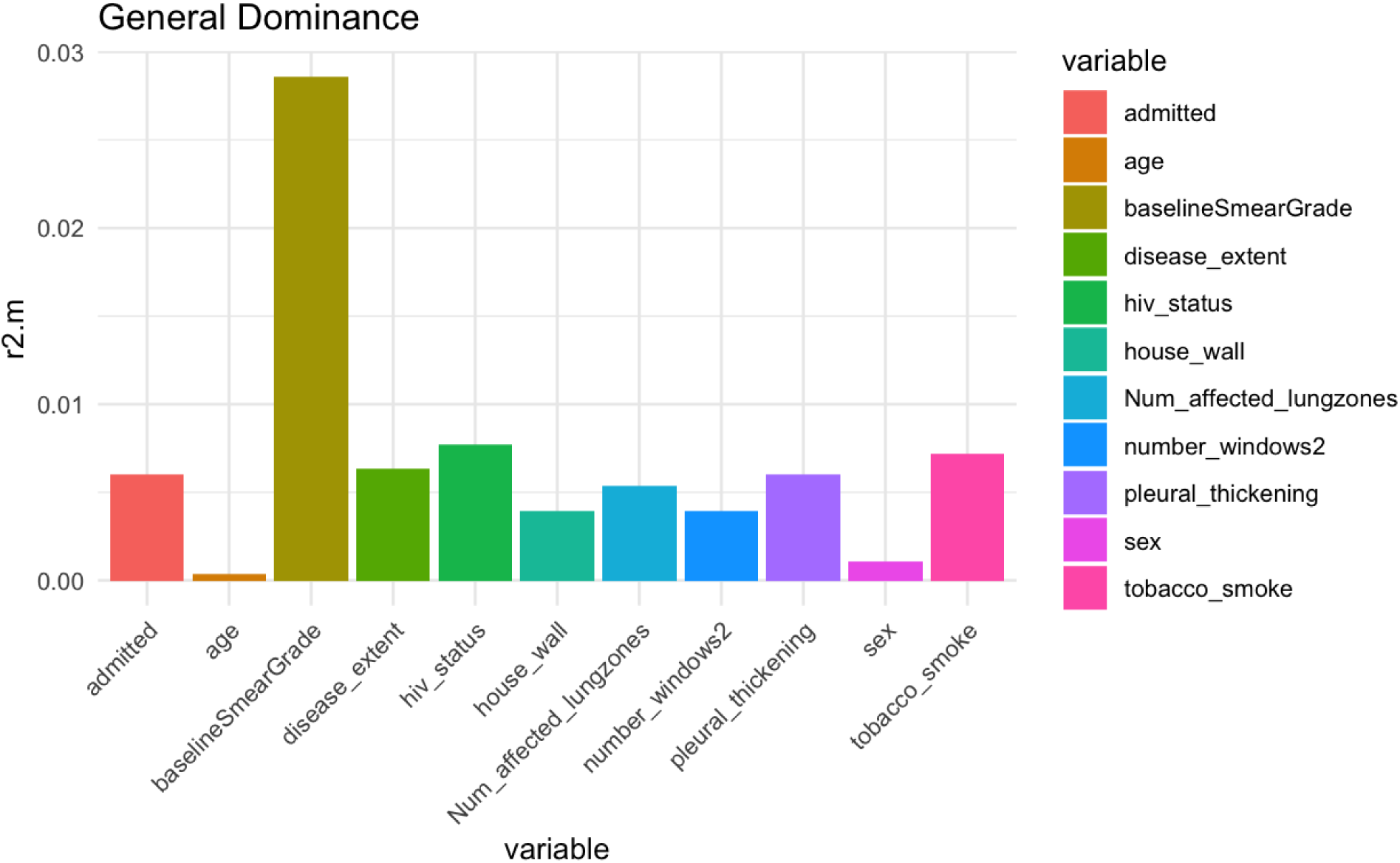
Dominance Analysis: (**a**) General dominance plot. The y-axis shows the fit statistics value (McFadden’s R-square); a measure of feature importance, the x-axis shows included features. (a) shows how the overall fit statistic value (McFadden’s R-square) is divided up among included features indicating that baseline smear grade generally dominates. (**b**) Conditional dominance plot. The y-axis shows the fit statistic (McFadden’s R-square) shows the McFadden’s R-square statistic associated with each feature related to the position (levels) at which it is included in the model, the x-axis shows levels/sequence at which features are included in the model. This shows that baseline smear also conditionally dominates other features.

**Figure 1b.**
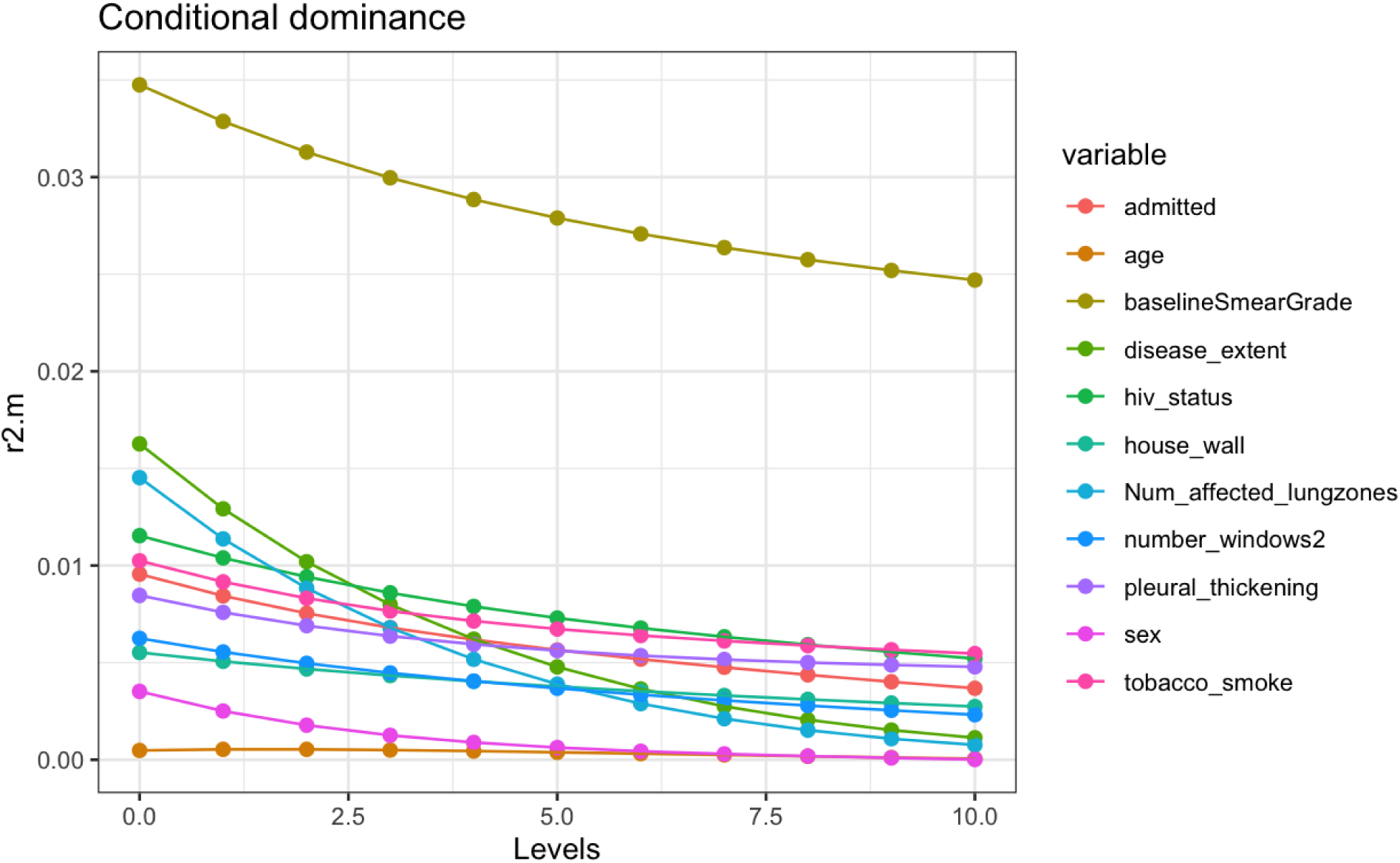
Conditional dominance

## Discussion

In high-burden and resource-limited settings, practical and accessible clinical tools that can identify individuals unlikely to culture convert are needed for risk stratification. Identifying factors that influence early culture conversion leads to better clinical monitoring guidelines. In this study, we assessed the association and predictive utility of two severity measures: the Bandim TBScore and CXR Score for culture conversion. We also identified contextual factors associated with culture conversion among adult index PTB cases enrolled in a community household study in Kawempe, Division of Kampala, Uganda. We found that most (approximately two-thirds) of cases culture converted, consistent with prior work[6, 10, 34]. We also found that the CXR Score and other features associated with culture conversion. However, although some measures of PTB severity did associate with conversion, none were highly predictive, highlighting that association does not always translate into prediction[35].

The finding that individuals in the highest CXR Score category, indicating the most severe disease, had lower odds of conversion, is consistent with prior reports associating CXR measures with poor conversion in drug sensitive[8, 36] and in multi drug-resistant TB (MDR-TB)[16, 37, 38] . However, unlike in a prior report[16], cavitations did not associate with culture conversion in our data even after stratifying by HIV status. Additionally, all participants in our cohort were drug sensitive cases. In contrast to prior studies that demonstrated association with treatment outcomes [18, 20, 39], the Bandim TBScore did not associate with culture conversion in our study. This is surprising as TBScore has associated with other clinical severity measures known to correlate with PTB outcomes in prior works[19, 36, 40]. Taken together, the differential associations between the CXR Score and TBScore with culture conversion indicate that these two measures operate at least semi-independently with respect to conversion or that each metric captures different domains of PTB severity.

The inverse association between baseline smear grade and culture conversion has been consistently reported in prior studies of both drug sensitive[3, 8, 9] and MDR-TB[6, 37]. Similarly, individuals presenting with pleural thickening were less likely to achieve culture conversion. Pleural thickening may result from pleural inflammation due to direct Mtb exposure or non-Mtb exposures such as asbestos[41], alcohol and tobacco smoking[42]. It reflects severe disease and may impair respiratory function[42, 43]. However, the mechanism by which it impedes conversion is less understood. Finally, current tobacco smokers were also less likely to convert, consistent with most prior work[5, 44, 45] even though some prior reports on this are inconsistent[46, 47].

The protective effect of HIV infection against poor culture conversion observed in our data seems counterintuitive considering that HIV and PTB synergistically exacerbate each other[48] and delays conversion especially in patients not on antiretroviral therapy (ART)[9, 10]. However, the relationship we found has previously been reported elsewhere[9]. HIV co-infected patients on care typically receive more intensive clinical follow-up, adherence counselling, prompt diagnosis and enrolment into care, all associated with better adherence and outcomes[9]. HIV co-infected patients tend to produce fewer amounts of Mtb (paucibacillary)[38, 49] and may be more easily colonized by multiple[50] less-fit Mtb strains that would not otherwise be pathogenic in immune competent individuals, potentially facilitating a more rapid conversion once TB treatment is initiated[51]. Therefore, the observed association likely reflects these influences rather than a direct protective effect of HIV co-infection.

While we found several variables that associated with culture conversion, our results indicate that these variables were limited in their prediction of culture conversion. The lack of predictive power of both severity measures, CXR Score and Bandim TBScore, is inconsistent with previous reports that found some severity measures predicted treatment outcomes and failures, but not explicitly culture conversion [8, 14, 18, 37]. However, the continued poor discriminative ability of associating variables in the most parsimonious model highlights an important need to identify additional and better predictors of early conversion in PTB care. Therefore, it might be useful to investigate a broader set of biomarkers, including genetic variants, for improved predictive value.

Dominance analysis indicated that baseline smear grade has the greatest overall contribution (general dominance), regardless of the position (level) at which it is included in the model with respect to competitor variables in the model (conditional dominance). This is an important finding that should inform future work given that it is not currently a component of either severity measure (CXR or TBscore). Therefore, future work to identify better predictors of culture conversion should include baseline smear grade, where possible. HIV status, tobacco smoking and disease extent were all conditionally dominant, indicating that their contribution varies depending on the position at which they are included in the model predicting culture conversion. Nonetheless, these should be considered in any models of culture conversion prediction.

We did not have data on treatment adherence and socio-economic status. These factors were therefore not accounted for in our models. However, treatment failure and death were rare in the parent study[52]. Given that these factors are strongly correlated with poor outcomes[6, 9] , it is unlikely that they biased our results. Also, our cohort was from a single urban site that may restrict generalizability. Finally, we recognize that individuals excluded from the study because they did not have culture conversion data were more likely to be thin, to have pleural effusion, anemia, disease in the left lung and to die. Hence, our analyses and conclusions may be more relevant to individuals with less severe disease.

Overall, our results indicate that prediction of conversion is not well captured by the numerous routinely collected variables we assessed despite some associations. This can impede effective clinical care. Hence, it is incumbent upon the field to identify a more useful set of variables and ones that can be used across high-risk environments beyond urban African settings.

## Data Availability

All data produced in the present study are available upon reasonable request to the authors

## Acknowledgements and Funding

Funding for this work was provided by the Tuberculosis Research Unit at the National Institutes of Health (grant N01-AI95383) and the National Institute of Allergy and Infectious Diseases (grants HHSN266200700022C/N01-AI70022, U19AI162583).

Funding for the first author (Lentlametse Mantshonyane) was provided by the Burroughs Wellcome Fund.

We thank the senior physicians, medical officers, health visitors, and laboratory and data personnel for their contributions: Dr. Lorna Nshuti, Dr. Roy Mugerwa, Dr. Alphonse Okwera, Dr. Deo Mulindwa, Denise Johnson, Dr. Allan Chiunda, Mark Breda, Dennis Dobbs, Mary Rutaro, Albert Muganda, Richard Bamuhimbisa, Yusuf Mulumba, Deborah Nsamba, Barbara Kyeyune, Faith Kintu, Gladys Mpalanyi, Janet Mukose, Grace Tumusiime, Pierre Peters, Annet Kawuma, Saidah Menya, Joan Nassuna, Keith Chervenak, Karen Morgan, Alfred Etwom, Micheal Angel Mugerwa, Emily Hellwig, and Lisa Kucharski. We also thank Dr. Francis Adatu Engwau, former Head of the Uganda National Tuberculosis and Leprosy Program, for supporting this project. We also thank the medical officers, nurses and counselors at the National Tuberculosis Treatment Centre, Mulago Hospital, the Ugandan National Tuberculosis and Leprosy Program and the Uganda Tuberculosis Investigation Bacteriological Unit, Wandegeya, for their contributions to this study.

## Conflict of interest

The Authors have no conflicts of interest to declare.

## Ethical Approval

This study was approved by National HIV/AIDS Research Committee of Makerere University and the institutional review board at University Hospitals, Cleveland Medical Center. Final clearance was given by the Uganda National Council for Science and Technology. Written informed consent was obtained from all study participants.

## Supplementary tables

**Supplementary Table 1.** Comparison of baseline characteristics between included cases and those excluded for lacking culture conversion data

| **Characteristic** | **Complete two-months culture conversion data<br>(Included cases,<br>N = 718)<sup>1</sup>** | **Missing two-months culture conversion data<br>(Excluded cases,<br>N = 157)<sup>1</sup>** | **p-value** |
|---|---|---|---|
| **Age(yrs)** | 29.26 (8.96) | 30.40 (9.49) | 0.15 |
| **Sex** |  |  | 0.49 |
| Female | 342 (48%) | 70 (45%) |  |
| Male | 376 (52%) | 87 (55%) |  |
| **HIV status** |  |  | 0.15 |
| Negative | 508 (71%) | 102 (65%) |  |
| Positive | 210 (29%) | 55 (35%) |  |
| **Marital status** |  |  | 0.51 |
| Married | 365 (51%) | 84 (54%) |  |
| Never married | 225 (31%) | 42 (27%) |  |
| Separated/divorced/widowed | 128 (18%) | 31 (20%) |  |
| **Smear grade at diagnosis** |  |  | 0.27 |
| Scanty | 22 (3.1%) | 9 (5.7%) |  |
| 1+ | 21 (2.9%) | 6 (3.8%) |  |
| 2+ | 85 (12%) | 21 (13%) |  |
| 3+ | 590 (82%) | 121 (77%) |  |
| **Tobacco smoking** |  |  | 0.47 |
| Never smoked | 589 (82%) | 123 (78%) |  |
| Former smoker | 116 (16%) | 30 (19%) |  |
| Currently smoke | 13 (1.8%) | 4 (2.5%) |  |
| **House lighting fuel** |  |  | 0.85 |
| Electricity | 293 (41%) | 68 (43%) |  |
| Kerosine | 400 (56%) | 84 (54%) |  |
| Other sources | 25 (3.5%) | 5 (3.2%) |  |
| **BandimTBScore** | 5.95 (2.10) | 6.15 (2.33) | 0.29 |
| **Weight Loss** | 654 (91%) | 142 (90%) | 0.80 |
| **Body Mass Index (Kg/m^2)** | 19.46 (2.81) | 19.27 (3.11) | 0.45 |
| **Fever (temp >37°C)** | 458 (64%) | 88 (56%) | 0.07 |
| **Tachycardia(pulse>90 beats/min)** | 561 (78%) | 130 (83%) | 0.19 |
| **Coughing** | 703 (98%) | 153 (97%) | 0.76 |
| **Hemoptysis** | 132 (18%) | 34 (22%) | 0.34 |
| **Night sweats** | 556 (77%) | 119 (76%) | 0.66 |
| **Dyspnea** | 311 (43%) | 71 (45%) | 0.66 |
| **Chest pains** | 495 (69%) | 98 (62%) | 0.11 |
| **Anemia** | 86 (12%) | 75 (48%) | <0.001 |
| **Lean Mass Index(kg/m^2)** | 16.19 (1.90) | 16.08 (1.71) | 0.50 |
| **Fat Mass Index(kg/m^2)** | 5.40 (4.85) | 4.29 (3.36) | 0.007 |
| **Positive findings auscultation** | 718 (100%) | 142 (100%) | 1.00 |
| **Chest X-ray Score** | 77.36 (37.99) | 72.51 (37.66) | 0.15 |
| **Extent of disease** |  |  | 0.71 |
| Normal | 21 (2.9%) | 6 (3.8%) |  |
| Minimal | 58 (8.1%) | 15 (9.6%) |  |
| Moderate | 250 (35%) | 57 (36%) |  |
| Advanced | 389 (54%) | 79 (50%) |  |
| **Affected Lung** |  |  | <0.001 |
| Both lungs | 375 (52%) | 51 (32%) |  |
| Right lung | 187 (26%) | 24 (15%) |  |
| Left lung | 156 (22%) | 82 (52%) |  |
| **Lung cavities** |  |  | 0.13 |
| No cavities | 264 (37%) | 68 (43%) |  |
| Cavities present | 454 (63%) | 89 (57%) |  |
| **Lung fibrosis** | 86 (12%) | 12 (7.6%) | 0.13 |
| **Lung adenopathy** | 34 (4.7%) | 8 (5.1%) | 0.84 |
| **Infiltrates observed** |  |  | 0.21 |
| No infiltrates | 29 (4.0%) | 6 (3.8%) |  |
| Lower lobe | 67 (9.3%) | 13 (8.3%) |  |
| Upper lobe | 113 (16%) | 31 (20%) |  |
| Both lobes | 509 (71%) | 107 (68%) |  |
| **Miliary disease** | 40 (5.6%) | 10 (6.4%) | 0.70 |
| **Pleural thickening** | 26 (3.6%) | 3 (1.9%) | 0.46 |
| **Pleural effusion** | 48 (6.7%) | 20 (13%) | 0.01 |
| **Number of affected lungs zones<sup>1</sup>** | 3 (2, 4) | 3 (2, 4) | 0.41 |
| **Admitted** | 72 (10%) | 18 (11%) | 0.59 |
| **Loss of appetite** | 481 (67%) | 101 (64%) | 0.52 |
| **Other respiratory diseases** | 58 (8.1%) | 10 (6.4%) | 0.47 |
| **Number of people per room<sup>1</sup>** | 4 (3, 6) | 4 (3, 6) | 0.30 |
| **Number of rooms<sup>1</sup>** | 1 (1, 3) | 1 (1, 2) | 0.34 |
| **Number of windows<sup>1</sup>** | 1 (0, 2) | 1(0, 2) | 0.49 |
| **Death** | 32 (4.5%) | 23 (14.6%) | <0.001 |

**Supplementary Table 2.** Correlation (r2>0.5) among variables significant at p<0.1 in Table 1

| **Variable1** | **Variable2** | **Correlation** |
|---|---|---|
| CXR_score | Num_affected_lungzones | 0.869 |
| CXR_score | Disease_extent | 0.761 |
| Disease_extent | Number_affected_lungzones | 0.704 |
| Disease_extent | Infiltrates | 0.691 |
| CXR_score | Infiltrates | 0.631 |
| Affected_Lung | Number_affected_lungzones | -0.629 |
| Infiltrates | Number_affected_lungzones | 0.623 |
| Affected_Lung | CXR_score | -0.566 |
| Muzingo | Number_windows | -0.565 |

**Supplementary Table 3.** Additional baseline characteristics considered for association with culture conversion

| **Characteristics** | **N (%)<br>Mean<br>(SD)** | **Two-month Culture conversion** |  | **p-value** |
|---|---|---|---|---|
|  |  | **Yes** | **No** |  |
| ***Other features*** | N=718 | 489 | 229 |  |
| Bandim TBScore |  |  |  |  |
| Weight loss (%) | 654(91.1) | 446 (91.2) | 208 (90.8) | 0.980 |
| Other resp disease (%) | 58 (8.1%) | 41 (8.4) | 17 (7.4) | 0.769 |
| Loss appetite (%) | 481(67%) | 321 (65.6) | 160 (69.9) | 0.300 |
| Alcohol = drinks alcohol (%) | 161(22%) | 114 (23.3) | 47 (20.5) | 0.460 |
| People per room (mean (SD)) | 3 (2, 4) | 2(2, 4) | 3 (2, 4) | 0.279 |
| Lung adenopathy (%) | 42 (5.8%) | 32 (6.5) | 10 (4.4) | 0.323 |
| Pleural effusion (%) | 48 (6.7%) | 30 (6.1) | 18 (7.9) | 0.482 |
| Miliary disease (%) | 33 (4.6%) | 23 (4.7) | 10 (4.4) | 0.992 |
| Biomass smoke (%) | 688 (96%) | 21 (4.3) | 9 (3.9) | 0.978 |
| House lighting fuel source (%) |  |  |  | 0.157 |
| Electricity | 468(65%) | 314 (64.2) | 154 (67.2) |  |
| Kerosine | 226(31%) | 162 (33.1) | 64 (27.9) |  |
| Other sources | 24(0.03%) | 13 (2.7) | 11 (4.8) |  |
| Coughing (%) | 703 (98%) | 478 (97.8) | 225 (98.3) | 0.874 |
| Hemoptysis (%) | 132 (18%) | 87 (17.8) | 45 (19.7) | 0.620 |
| Fever at baseline (temp >37°C) | 458 (64%) | 310 (63.4) | 148 (64.6) | 0.812 |
| Night sweats (%) | 556(77%) | 383 (78.3) | 173 (75.5) | 0.463 |
| Dyspnea (%) | 311 (43%) | 206 (42.1) | 105 (45.9) | 0.391 |
| Positive finding at ausc = yes (%) | 694 (97%) | 469 (95.9) | 225 (98.3) | 0.160 |
| Chest pains = Yes (%) | 495 (69%) | 345 (70.6) | 150 (65.5) | 0.202 |
| Anemia | 86 (12%) | 61 (12.5) | 25 (10.9) | 0.634 |
| Lung cavities (%) | 460 (64%) | 305 (62.4) | 155 (67.7) | 0.194 |

**Supplementary Table 4.** Univariate Associations between components of the Bandim TBScore and culture conversion

| **Bandim components** | **OR (95% CI)** | **p.value** |
|---|---|---|
| Coughing: Yes | 0.77 (0.21, 2.29) | 0.66 |
| Fever: Yes | 0.96 (0.69, 1.32) | 0.79 |
| Chest pains: Yes | 1.26 (0.90, 1.76) | 0.17 |
| Night sweats: Yes | 1.17 (0.80, 1.69) | 0.41 |
| Dyspnea: Yes | 0.85 (0.61, 1.17) | 0.32 |
| Anemia: Yes | 1.10 (0.71, 1.74) | 0.68 |
| Tachycardia: Yes | 1.43 (0.99, 2.07) | 0.06 |
| Low lean mass index: Yes | 0.92 (0.65, 1.31) | 0.65 |
| Low fat mass index: Yes | 1.18 (0.85, 1.65) | 0.32 |
| BMI; Low(<18kg/m2) | 1.00 (0.73, 1.37) | 0.99 |
| Hemoptysis: Yes | 0.88 (0.60, 1.33) | 0.55 |
| Positive auscultation: Yes | 0.80 (0.50, 1.26) | 0.34 |

**Supplementary Table 5.** Dominance Analysis: Average contributions by Predictor based on 4 different fit indices of the logistic regression model

| Average Contribution by predictor |  |  |  |  |  |  |  |  |  |  |  |
|---|---|---|---|---|---|---|---|---|---|---|---|
| Fit Index | Baseline Smear grade | Pleural thickening | Admitted | Tobacco smoke | Disease extent | Number affected lung zones | Number of windows | House wall | Age | Sex | HIV status |
| r2.m | 0.029 | 0.006 | 0.006 | 0.007 | 0.006 | 0.005 | 0.004 | 0.004 | 0 | 0.001 | 0.008 |
| r2.cs | 0.034 | 0.007 | 0.007 | 0.009 | 0.008 | 0.006 | 0.005 | 0.005 | 0 | 0.001 | 0.009 |
| r2.n | 0.048 | 0.010 | 0.010 | 0.012 | 0.011 | 0.009 | 0.007 | 0.007 | 0.001 | 0.002 | 0.013 |
| r2.e | 0.035 | 0.007 | 0.007 | 0.009 | 0.008 | 0.007 | 0.005 | 0.005 | 0 | 0.001 | 0.010 |

\* Fit Indices: r2.m (McFadden’s measure), r2.cs (Cox and Snell’s measure), r2.n (Nagelkerke’s measure), r2.e (Estrella’s measure)

## Notes

### Competing Interest Statement

The authors have declared no competing interest.

